# Sustained Human Outbreak of a New MPXV Clade I Lineage in Eastern Democratic Republic of the Congo

**DOI:** 10.1101/2024.04.12.24305195

**Authors:** Emmanuel H. Vakaniaki, Cris Kacita, Eddy Kinganda-Lusamaki, Áine O’Toole, Tony Wawina-Bokalanga, Daniel Mukadi-Bamuleka, Adrienne Amuri Aziza, Nadine Malyamungu-Bubala, Franklin Mweshi Kumbana, Léandre Mutimbwa-Mambo, Freddy Belesi-Siangoli, Yves Mujula, Edyth Parker, Pauline-Chloé Muswamba-Kayembe, Sabin S. Nundu, Robert S. Lushima, Jean Claude Makangara Cigolo, Noella Mulopo-Mukanya, Elisabeth Pukuta Simbu, Prince Akil-Bandali, Hugo Kavunga, Koen Vercauteren, Nadia A. Sam-Agudu, Edward J Mills, Olivier Tshiani-Mbaya, Nicole A. Hoff, Anne W. Rimoin, Lisa E. Hensley, Jason Kindrachuk, Ahidjo Ayouba, Martine Peeters, Eric Delaporte, Steve Ahuka-Mundeke, Jean B. Nachega, Jean-Jacques Muyembe-Tamfum, Andrew Rambaut, Laurens Liesenborghs, Placide Mbala-Kingebeni, the Mpox Research Consortium

**Affiliations:** Institut National de Recherche Biomédicale, Kinshasa, Democratic Republic of Congo; Hemorrhagic Fever and Monkeypox Program, Ministry of Health, Kinshasa, Democratic Republic of the Congo; Service de Microbiologie, Département de Biologie médicale, Cliniques Universitaires de Kinshasa, Université de Kinshasa; TransVIHMI (Recherches Translationnelles sur le VIH et les Maladies Infectieuses endémiques et émergentes), Université de Montpellier, French National Research Institute for Substaminale Développent (IRD), INSERM, Montpellier, France; Institute of Ecology and Evolution, University of Edinburgh, Edinburgh EH9 3FL, UK; Department of Clinical Sciences, Institute of Tropical Medicine, Antwerp, Belgium; Rodolphe Merieux INRB-Goma Laboratory, Goma, Democratic Republic of the Congo; Kamituga General Hospital, South Kivu, Democratic Republic of Congo; Kamituga Health Zone, South Kivu, Democratic Republic of Congo; Provincial Health Division, South Kivu, Democratic Republic of Congo; African Center of Excellence for Genomics of Infectious Diseases, Redeemer’s University, Ede, Osun State, Nigeria; Department of Pediatrics and Child Health, School of Medical Sciences, University of Cape Coast, Cape Coast, Ghana; International Research Center of Excellence, Institute of Human Virology Nigeria, Abuja, Nigeria; Global Pediatrics Program and Division of Infectious Diseases, Department of Pediatrics, University of Minnesota Medical School, Minneapolis, MN, USA; Department of Health Research Methods, Evidence and Impact, Faculty of Health Sciences, McMaster University, Hamilton, Ontario, Canada; Frederick National Laboratory, Leidos Biomedical Research, Clinical Monitoring Research Program Directorate; Department of Epidemiology, Jonathan and Karin Fielding School of Public Health, University of California, Los Angeles, CA, USA; US Department of Agriculture, Manhattan, KS, USA; University of Manitoba, Winnipeg, Manitoba, Canada; Department of Epidemiology, Infectious Diseases and Microbiology, University of Pittsburgh, School of Public Health, Pittsburgh, PA, USA; Department of Epidemiology and International Health, Johns Hopkins Bloomberg School of Public Health, Baltimore, MD, USA; Department of Medicine, Division of Infectious Diseases, Stellenbosch University Faculty of Medicine and Health Sciences, Cape Town, South Africa; Department of Microbiology, Immunology and Transplantation, KU Leuven, Leuven, Belgium

## Abstract

**Background:** Monkeypox virus (MPXV) attracted global attention in 2022 during a widespread outbreak linked primarily to sexual contact. Clade I MPXV is prevalent in Central Africa and characterized by severe disease and high mortality, while Clade II is confined to West Africa and associated with milder illness. A Clade IIb MPXV emerged in Nigeria in 2017, with protracted human-to-human transmission a forerunner of the global Clade II B.1 lineage outbreak in 2022. In October 2023, a large mpox outbreak emerged in the Kamituga mining region of the Democratic Republic of the Congo (DRC), of which we conducted an outbreak investigation.

**Methods:** Surveillance data and hospital records were collected between October 2023 and January 2024. Blood samples and skin/oropharyngeal swabs were obtained for molecular diagnosis at the National Institute of Biomedical Research, Kinshasa. MPXV genomes were sequenced and analyzed using Illumina NextSeq 2000 and bioinformatic tools.

**Results:** The Kamituga mpox outbreak spread rapidly, with 241 suspected cases reported within 5 months of the first reported case. Of 108 confirmed cases, 29% were sex workers, highlighting sexual contact as a key mode of infection. Genomic analysis revealed a distinct MPXV Clade Ib lineage, divergent from previously sequenced Clade I strains in DRC. Predominance of APOBEC3-type mutations and estimated time of emergence around mid-September 2023 suggest recent human-to-human transmission.

**Conclusions:** Urgent measures, including reinforced, expanded surveillance, contact tracing, case management support, and targeted vaccination are needed to contain this new pandemic-potential Clade Ib outbreak.

## Introduction

Monkeypox virus (MPXV) attracted global attention in 2022 due to a widespread outbreak linked to sexual contact. For more than 40 years prior, MPXV was responsible for an increasing number of outbreaks in several endemic African countries, predominantly from zoonotic spillover. ^1^ MPXV is an enveloped, doublelJstranded DNA virus of the *Poxviridae* family that includes vaccinia, cowpox, smallpox, and several pathogenic animal viruses. In humans, the virus causes mpox, a disease characterized by fever, lymphadenopathy, and a vesiculo-papular rash. Two distinct genetic MPXV clades are known: Clade I is predominantly found in Central Africa, particularly in the Democratic Republic of the Congo (DRC), and is characterized by severe clinical symptoms and high mortality of up to 11%.^2^ Conversely, Clade II, confined to West Africa until the 2022 global epidemic, causes less severe illness and lower mortality of <4%.^2^

Historically, Clade I MPXV has predominated, accounting for 95% of reported cases, while Clade II infections have been less common.^1^ This changed in 2017 when a large outbreak of Clade IIb MPXV occurred in Nigeria, with indications of sustained human-to-human transmission, including through sexual contact.^3^ These findings were largely overlooked until a related Clade IIb lineage —designated B.1^4^— spawned a global outbreak in May 2022, with 94,707 confirmed cases in 117 countries as of February 2024. ^4^ Genomic analyses of the B.1 lineage revealed a skewed mutational pattern suggestive of non-canonical evolution, likely driven by an apolipoprotein B mRNA editing enzyme, catalytic subunit 3 (APOBEC3) cytosine deamination, a characteristic feature of human MPXV infection.^5,6^ MPXV mutations have been confirmed *in vitro* as due to the action of APOBEC3F, suggesting that Clade IIb may have entered human populations as early as 2015. ^7,8^ As of March 2024, the global Clade IIb B.1 epidemic has largely subsided, although small outbreaks still occur in Nigeria and other endemic countries.^4^

As Clade II outbreaks have waned, Clade I MPXV infections in Central Africa have been increasing,^4^ especially in remote forest areas, likely due to zoonotic spillover. There is limited documentation of secondary human-to-human transmission within households.^9,10^ Nevertheless, progressive annual increases in mpox cases reported in the DRC, with a record 14,626 suspected cases in 2023, suggest a shift toward increased human-to-human transmission.^11^ We recently documented a cluster of MPXV infections caused by sexual contact in the DRC, although the transmission chain was not sustained.^12^ Concurrently, new mpox cases have continued to emerge in several previously unaffected areas of the DRC. In October 2023, the first-ever mpox cases were detected in Kamituga Health Zone, a densely populated mining area in South Kivu province in Eastern DRC. In this report, we describe the results of a recent outbreak investigation in this area.

## Methods

### Population and setting

This study included patients hospitalized at the General Hospital of Kamituga, a gold mining town with approximately 242,000 inhabitants (**Figure 1A**).

**Figure 1.**
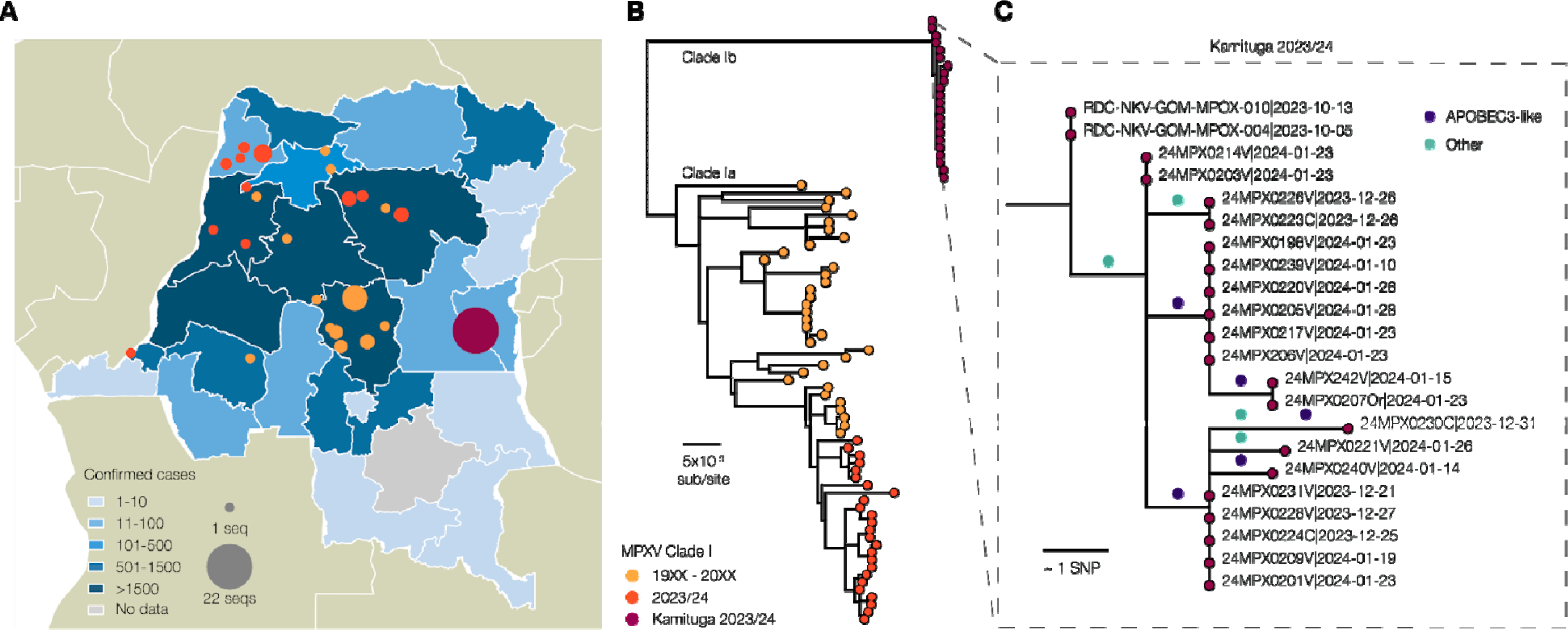
**A)** Map of the Democratic Republic of Congo with Provinces colored by the number of reported cases. MPXV genomes available are indicated by colored circles. **B)** Maximum likelihood phylogeny of Clade I genomes, including those from the present study (indicated as 2023/24 and Kamituga samples), with all publicly available previously sequenced genomes. A labelled version of this phylogeny can be found in the supplementary materials (Supplementary Figure 1). **C)** The Kamituga cluster with reconstructed mutations indicated on the branches. Mutations are colored by whether they are consistent with APOBEC3 deamination or not.

## Sampling and Data Collection

A multidisciplinary team including representatives from the Ministry of Health, provincial health authorities, and the National Institute for Biomedical Research (INRB) investigated the mpox outbreak in Kamituga in January 2024. We collated mpox monitoring data collected by provincial surveillance authorities. Local surveillance teams investigated and collected data from each suspected mpox case using a standardized Ministry of Health form. This form contains questions on demographics (age, sex, residence, profession, nationality), clinical symptoms, outcome, and mpox sampling. For additional clinical data, we reviewed medical records of patients with suspected mpox admitted to the hospital between October 6, 2023, and January 23, 2024. We then conducted interviews with local healthcare workers (HCWs) and local provincial and municipal health authorities. Our INRB team interviewed and examined patients with suspected mpox, collecting samples from blood, skin lesions, and oropharyngeal swabs for molecular diagnosis. Real-time PCR assays were performed at INRB Goma using the LightMix Modular Monkeypox virus kit (Roche, Germany) according to manufacturer’s instructions.

## MPXV Genome Sequencing

Sequencing was performed at the INRB’s Pathogen Genomics lab in Kinshasa. Samples testing positive with Cycle threshold (Ct) values <31 underwent sequencing using Illumina DNA Prep with Enrichment kit (Illumina) and Comprehensive Viral Research Panel (Twist Biosciences). Following manufacturer’s protocol, libraries were prepared and loaded onto the NextSeq 2000 sequencer. FASTQ files were processed through GeVarLi (https://forge.ird.fr/transvihmi/nfernandez/GeVarLi), CZid (https://czid.org/) and iVar pipelines for read quality control, consensus genome coverage, and variant calling using a Clade I reference genome (an early MPXV genome from DRC, accession: NC_003310).

## Phylogenetic analysis

We compiled a dataset including all high-quality Clade I MPXV genome sequences from Genbank (accessions and author list in **Supplementary Table 1**) and used Clade IIa MPXV genomes from Nigeria dated 1978 and 1971 as outgroups (accession numbers KJ642615 and KJ642617, respectively). We estimated a maximum likelihood phylogeny using IQ-TREE 2 version 2.2.5 with HKY substitution model. ^13,14^ We also analyzed genome fragments from three mpox cases from North and South Kivu in 2011-2012 ^15^ (**Supplemental Figure 1**). Ancestral reconstruction was performed for each internal node on the phylogeny using IQ-TREE 2, enabling mapping of single nucleotide polymorphisms (SNPs) along branches. SNPs were categorized based on whether they were consistent with the signature of APOBEC3-editing, assuming this process induced specific mutations (TC➔TT and GA➔AA). We conducted a comparative analysis of isolates from Kamituga against viruses recently isolated in concurrent outbreaks in several DRC provinces. We used O’Toole’s method ^7^ to estimate the date of the most recent common ancestor of the Kamituga MPXV genomes and establish timing for outbreak emergence. This approach involves partitioning the alignment into segments containing sites potentially subject and not subject to APOBEC3 editing. We constrained the rate of evolution of these two partitions using posterior estimates from O’Toole (approximated by a normal with mean 1.3 x 10^4^ substitutions/site/year and standard deviation 1.0 x 10^4^; and mean 4.2 x 10^6^ and standard deviation 0.49x10^6^, respectively). We employed an exponential growth coalescent model as a prior on the tree.

## Research Ethics Committee Approval

This study used anonymized data from mpox surveillance activities, outbreak response surveys, and retrospective medical chart review and was approved by the Ethics Research Committee of the University of Kinshasa School of Public Health (ESP/CE/78/2024).

## Data Availability

All epidemiological data produced in the present study are available upon reasonable request to the authors. Sequencing data are publicly accessible (https://github.com/inrb-labgenpath/Mpox_sequencing_Kamituga).

## Results

A total of 241 suspected mpox cases were recorded by South Kivu’s provincial surveillance between September 29, 2023, and February 29, 2024 (**Figure 2A**). Most cases (93%) were reported in Kamituga Health Zone, and most (91%) were hospitalized. Samples from 119 (48.9%) individuals were collected for PCR testing at INRB, Goma, and108/119 suspects (90.8%) were confirmed MPXV-positive. Demographic characteristics and clinical manifestations were similar between suspected and confirmed cases (**Table 1**).

**Figure 2.**
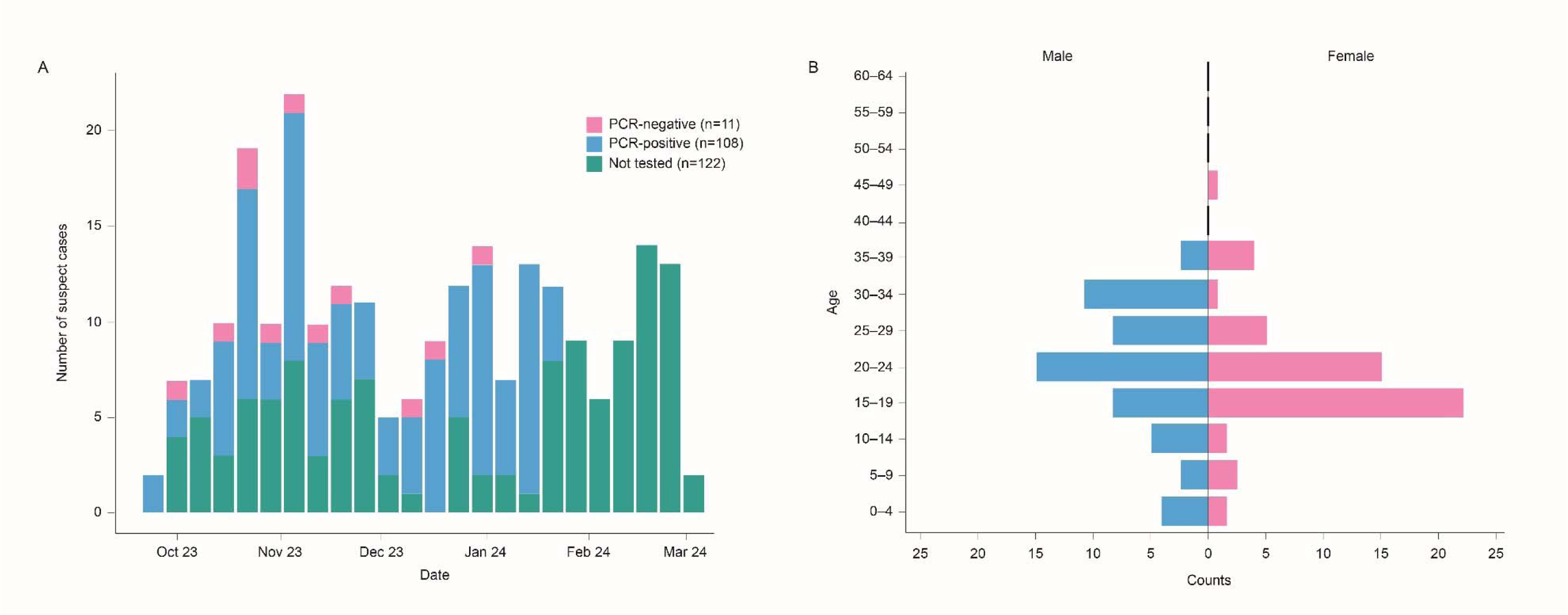
**A)** Epidemiologic curve of Kamituga mpox outbreak (October 2023 to March 2024); **B**) Counts of suspected mpox cases disaggregated by age and sex (Male in blue; Female in pink)

**Table 1:** Clinical and demographic characteristics of mpox suspect and confirmed cases.

|  |  |  |
| --- | --- | --- |
| **Characteristic** | **Suspected cases**<br>**n = 241** | **Confirmed cases**<br>**n = 108** |
| Health zone where the diagnosis was made – no. (%) |  |  |
| Kamituga | 224 (93%) | 103 (95%) |
| Other | 17 (7.1%) | 5 (4.6%) |
| Age (years) – median (IQR) | 21 (18, 28) | 22 (18, 27) |
| Sex – no. (%) |  |  |
| Female | 128 (54%) | 56 (52%) |
| Male | 111 (46%) | 52 (48%) |
| Unknown | 2 | 0 |
| Professional sex worker – no. (%) | 71 (29.5%) | 31 (28.7% %) |
| Smallpox vaccinated – no. (%) |  |  |
| No | 239 (100%) | 108 (100%) |
| Unknown | 2 | 0 |
| Nationality – no. (%) |  |  |
| Congolese | 217 (99%) | 92 (99%) |
| Rwandese | 1 (0.5%) | 1 (1.1%) |
| Burundese | 2 (0.9%) | 0 (0%) |
| Unknown | 21 | 15 |
| Hospitalized – no. (%) |  |  |
| Yes | 217 (91%) | 104 (96%) |
| No | 22 (9.2%) | 4 (3.7%) |
| Unknown | 2 |  |
| Bedridden – no. (%) |  |  |
| Yes | 9 (3.8%) | 10 (9.3%) |
| No | 224 (95%) | 98 (91%) |
| Unknown | 3 | 0 |
| Fever at presentation – no. (%) |  |  |
| Yes | 148 (62%) | 64 (59%) |
| No | 91 (38%) | 44 (41%) |
| Unknown | 2 | 0 (0%) |
| Skin rash at presentation – no. (%) |  |  |
| Yes | 239 (100%) | 108 (100%) |
| No | 0 (0%) | 0 (0%) |
| Unknown | 0 | 0 |
| Cough at presentation – no. (%) |  |  |
| Yes | 41 (18%) | 20 (19%) |
| No | 191 (82%) | 85 (81%) |
| Unknown | 9 | 3 |
| Adenopathy at presentation – no. (%) |  |  |
| Yes | 99 (42%) | 45 (42%) |
| No | 138 (58%) | 62 (58%) |
| Unknown | 4 | 1 |
| Throat ache at presentation – no. (%) |  |  |
| Yes | 57 (24%) | 29 (27%) |
| No | 180 (76%) | 78 (73%) |
| Unknown | 4 | 1 |
| Oral ulcers at presentation – no. (%) |  |  |
| Yes | 24 (10%) | 13 (12%) |
| No | 213 (90%) | 94 (88%) |
| Unknown | 4 | 1 |
| Headache at presentation – no. (%) |  |  |
| Yes | 43 (18%) | 19 (18%) |
| No | 194 (82%) | 88 (82%) |
| Unknown | 4 | 1 |
| Myalgia at presentation – no. (%) |  |  |
| Yes | 41 (17%) | 20 (19%) |
| No | 196 (83%) | 87 (81%) |
| Unknown | 4 | 1 |
| Fatigue at presentation – no. (%) |  |  |
| Yes | 30 (13%) | 19 (18%) |
| No | 207 (87%) | 88 (82%) |
| Unknown | 4 | 1 |
| Conjunctivitis at presentation – no. (%) |  |  |
| Yes | 9 (3.8%) | 8 (7.4%) |
| No | 229 (96%) | 100 (93%) |
| Unknown | 3 | 0 |

Most individuals with confirmed MPXV infection were female (56/108, 51.9%). The median age was 22 years (IQR 18-27) (**Figure 2B**). Children under 15 years constituted 14.8% (16/108) of confirmed cases, individuals aged 15-30 years accounted for 67% (73/108), and those 30-49 years comprised 17.6% (19/108) of cases. Additionally, 28.7% (31/108) of confirmed cases and 29.5% (71/241) of all suspected cases occurred among professional sex workers from Kamituga. None of the individuals with confirmed mpox had been vaccinated against smallpox.

The provincial survey form included a limited number of clinical variables. All confirmed cases presented with a cutaneous rash, 64/108 (59%) presented with fever, and 45/107 (42%) with adenopathy. Most mpox-suspected individuals underwent hospital admission, but this was mostly for isolation purposes. Among confirmed cases, 10/108 (9.3%) were bedridden (**Table 1**). No deaths were recorded in the provincial database. To obtain more information on clinical presentation, we reviewed hospital records and identified 148 patients admitted with suspected mpox, and additional clinical information was available for 134 of these patients. HIV status was known for 31% (46/148) of these patients, with 6.5% (3/46) reportedly positive. Eighty-five percent (114 patients) presented with genital lesions. Interviews conducted with 25 individuals with confirmed mpox revealed that 22 had physical contact with another MPXV-infected patient, 13 of whom specifically reported sexual contact. Two mpox patients (1.4%) died during hospitalization.

Near full-length MPXV genome sequences, including 12 with genome coverage above 99%, three between 95% and 99%, and two between 90% and 95%, were obtained from 22 patients. Genomic analysis revealed that all MPXV strains isolated from Kamituga clustered tightly with each other on a distinct lineage from previously sequenced Clade I MPVX (**Figure 1**). The new lineage increases the known diversity of Clade I by an additional 54% (**Supplemental Figure 2**). Within the clade that previously encompassed the entirety of Clade I, the maximum pairwise patristic distance totaled 4.89 x 10^4^ substitutions per site (equivalent to ∼96 individual nucleotide changes). With the inclusion of genomes from the Kamituga outbreak, this increased to 7.55 x 10^4^ substitutions per site, or ∼149 nucleotide changes. Sequences from two 2011-2012 MPXV samples from South Kivu and North Kivu provinces^15^ clustered with the genomes sequenced in this study (**Supplemental Figure 1).** However, due to limited genome coverage, their precise phylogenetic position relative to the Kamituga cluster could not be reliably determined. These patients were from rural areas and were temporally and geographically distinct, with no epidemiological linkage. Therefore, it is likely that these cases arose from zoonotic spillover, suggesting that this lineage is endemic in regionally based animal reservoir(s).

The 22 MPXV genome sequences from Kamituga formed a compact cluster with a total of nine reconstructed mutations (**Figure 1B and 1C**). Among these, five were consistent with APOBEC3-mediated cytosine deamination, indicative of human-to-human transmission. Based on the observed frequency of APOBEC3-like mutations in Clade I MPXV in reservoir host species (8%)^7^, one would expect fewer than one APOBEC3-like mutation out of a total nine mutations.

Furthermore, if MPXV in this cluster continued to evolve as observed in rodent host species, the binomial probability of observing five or more APOBEC3 mutations, would be 0.0003. The observed predominance of APOBEC3-type mutations supports the finding that this entire cluster resulted from human-human transmission, whereas the limited overall genomic diversity indicates that this outbreak began recently.

Molecular clock analysis, using the rate of evolution estimated for human Clade IIb outbreak ^7^, estimates that the most recent common ancestor of the Kamituga genomes existed around mid-September 2023, which is consistent with the earliest reported cases. However, the credible intervals of this estimate cannot rule out a date as early as July 2023 (mean estimate 13-Sep-2023 with 95% highest posterior density intervals of 9-Jul-2023 to 3-Oct-2023). The estimated exponential growth rate was 10.8 per year (doubling time of 23 days), but the credible 95% density intervals include zero.

The remaining genome sequences from this study, i.e., those sampled from cases in other provinces from late 2023 to early 2024 (**Figure 1B**), aligned with previous Clade I diversity (Clade Ia), indicating no connection to the Kamituga outbreak (**Figure 1B**). Furthermore, the proportion of APOBEC3-type mutations compared to other mutations in samples collected outside Kamituga province was only 15.3% (15 of 98 observed mutations), suggesting limited human-to-human transmission in those areas.

## Discussion

This report describes a novel Clade I MPXV lineage associated with sustained human-to-human transmission in an ongoing outbreak in eastern DRC. Identification of APOBEC3-related mutations – the hallmark of efficient MPXV spread via human-to-human transmission – bolsters this assertion. Due to its distinct geographical location and divergent phylogenetic relationship, we propose to name this new Clade Ib, with the previously described Clade I renamed Clade Ia. The previous distinction between Clades IIa and IIb was borne out of a similar rationale, with the diversity within Clades IIa and IIb more than half the divergence between them. ^16^ Although the proposed Clade Ib is currently only represented by genomes from the Kamituga outbreak, the short fragmentary sequences isolated from earlier zoonotic mpox cases in the area suggest that this lineage likely pre-existed in a local, non-human animal reservoir. ^15^

Of note, a large ∼1kbp deletion in the MPXV genome (Δ19,128-20,270 coordinates relative to the Clade I reference genome, GenBank accession: NC_003310) has been reported in genome sequences from Kamituga linked to this outbreak, ^14^ interfering with the Clade I-specific diagnostic PCR originally developed by Li *et al.* ^17^ The new genomes associated with the Kamituga outbreak also contain this deletion, implying that all genomes sampled from Clade Ib thus far exhibit this deletion and will not amplify with the specific diagnostic. Conversely, the other MPXV Clade Ia genomes we sampled from 2023/2024 did not contain this deletion. The two mpox samples from a 2011-2012 outbreak in the region^15^ clustered with the Kamituga outbreak genomes, but only had five gene fragments sequenced with no coverage of the deleted region. We cannot say whether this is a recent deletion specific to this outbreak or whether it was a viral feature in a non-human animal reservoir for this lineage.

Strengths of our study include novel and high-quality viral genomic sequencing performed at quality-controlled national and international laboratories in endemic mpox regions. However, limitations include the limited availability of comprehensive clinical and demographic patient information in this retrospective case series. Despite these constraints, our data suggest that transmission in this outbreak was primarily linked to sexual contact: first, most affected individuals were adolescents and young adults, contrasting with previous mpox outbreaks in the DRC, where children under 15 years were most affected; ^18,19^ second, professional sex workers were disproportionately affected; finally, hospital records indicate that most suspected cases presented virus-compatible genital lesions.

The sustained spread of clade I MPXV in Kamituga, a densely populated, poor mining region, raises significant concerns. Local healthcare infrastructure is ill-equipped to handle a large-scale epidemic, compounded by limited access to external aid. The 241 reported cases are likely an underestimate of the true prevalence of mpox cases occurring in the area. In conversations with local HCWs, they reported that many people with mpox signs and symptoms remain in the community and do not seek care.

Frequent travel occurs between Kamituga and the nearby city of Bukavu, with subsequent movement to neighboring countries such as Rwanda and Burundi. Moreover, a considerable number of sex workers operating in Kamituga are foreigners and frequently return to their countries of origin, although at present, there is no evidence of wider dissemination of the outbreak. The highly mobile nature of this mining population poses a substantial risk of outbreak escalation beyond the current area and across borders.

Our data supports a signature of human-to-human transmission in MPXV genomes from the Kamituga region, supporting epidemiological investigations that suggested transmission through sexual contact in affected locations. However, this cluster of patients in Kamituga represents a minority of mpox cases reported during the 2022/2023 MPXV surge in the DRC. Genomic evidence demonstrates that most cases are not due to a single human outbreak or increased transmissibility from virus changes. Instead, these cases are likely due to several independent spillover events from reservoir hosts. It is unclear why this surge in Kamituga cases has occurred. Further investigation is needed to understand the drivers of these spillover events.

The situation in Kamituga echoes the 2017-2018 outbreak of Clade IIb MPXV in Nigeria. Without intervention, this localized Kamituga outbreak harbors the potential to spread nationally and internationally. Urgent measures must be implemented, including intensifying local surveillance, enhancing referral systems and case management, and implementing targeted mpox vaccination for individuals at high-risk, such as contacts of index cases, HCWs, sex workers, and men who have sex with men. ^20^ Given the recent history of mpox outbreaks in DRC, we advocate for swift action by endemic countries and the international community to avert another global mpox outbreak.

## Data Availability

All epidemiological data produced in the present study are available upon reasonable request to the authors. Sequencing data are available on the INRB Github page

https://github.com/inrb-labgenpath/Mpox_sequencing_Kamituga.git

## Acknowledgments

We thank John L. Johnson, MD, Case Western Reserve University, CL, OH, USA; Philip J Rosenthal, MD, University of California San Francisco, CA, USA; Michel P. Hermans, MD, PhD, Université Catholique Louvain, Brussels, Belgium; and Wolfgang Preiser, MD, PhD, Stellenbosch University, Cape Town, South Africa; and Lee Harrison, MD, University of Pittsburgh, PA, USA for critical review of the manuscript. We are sincerely grateful to all Kamituga staff involved in the management of this outbreak, the health division province in South Kivu, and the Ministry of Health. Thanks also to the clinical and provincial staff in Kamituga (Léandre Mutimbwa-Mambo, Nadine Malyamungu-Bubala, Freddy Belesi-Siangoli, Franklin Mweshi-Kumbana, Ombotimbe Abdramane, Justin Bengehya Mbiribindi, Gaston Lubambo, MD, Willy Kasi) and the logistics team in Goma that helped transfer the samples to Kinshasa. We also acknowledge the support of all the INRB epidemiology, Virology and Biorepository team (Elise Muyamuna, Sophie Gabia, Andre Citenga, Francisca Muyembe, Ola Rilia, Gradi Luakanda, Emmanuel Lokilo, Princesse Paku and Sifa Kavira) and Microbiology staff (Michel Kenye, Raphael Lumembe and Gabriel Kabamba). We also thank all members of the MpoxReC (see supplemental Table 1) as well as members of the Africa Pathogen Genomics Initiative (CARES), AFROSCREEN, DTRA, USDA, and IMReC for supporting the outbreak response and the genomic surveillance in DRC.

---

**Supplemental Figure 1.**
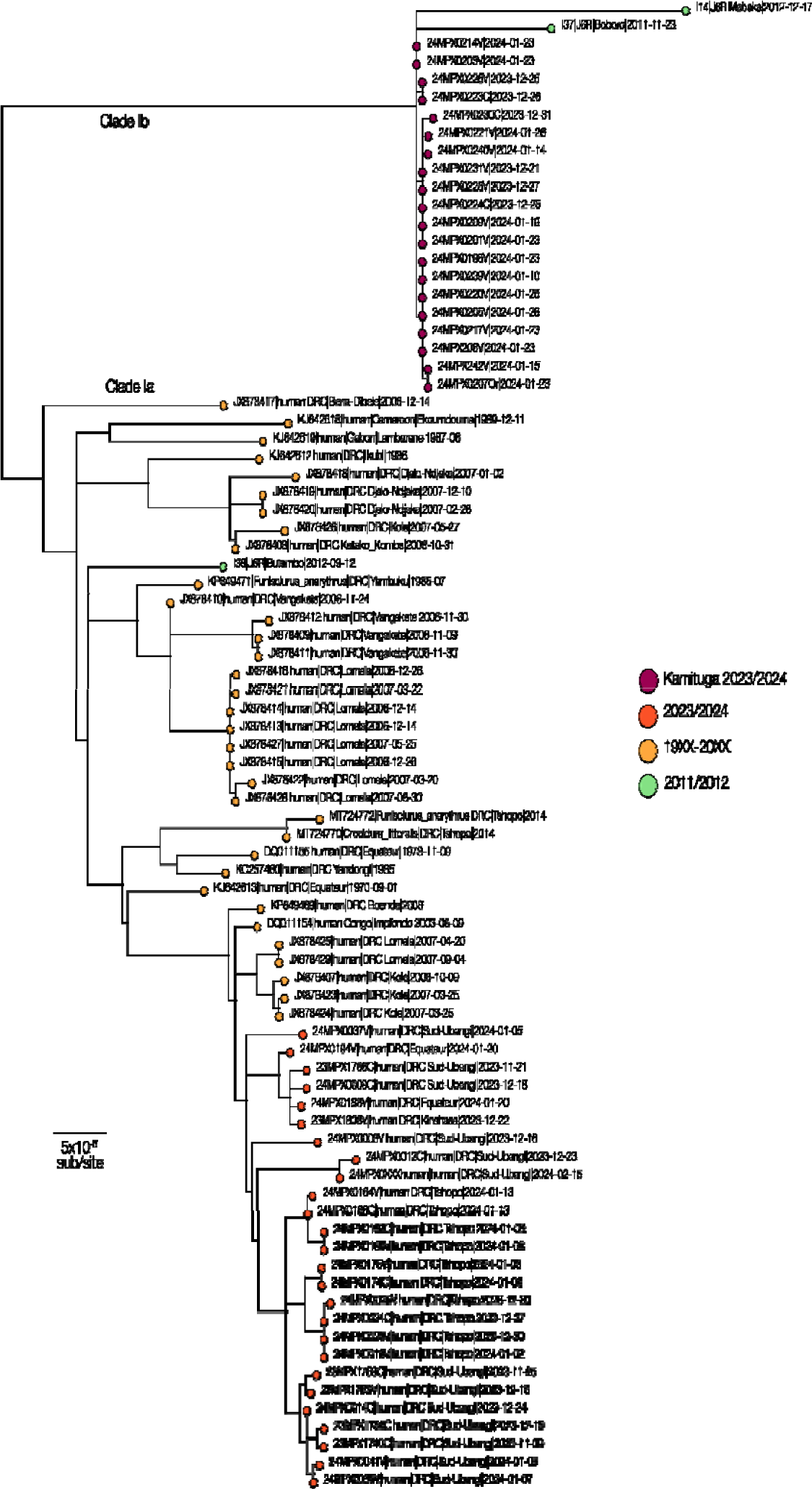
We estimated a labeled maximum likelihood phylogeny using IQTREE2, including all high quality MPXV Clade I genome sequences available on Genbank. We also include a set of sequences constructed from fragments sequenced from an outbreak in Sud-Kivu and Nord-Kivu, reported in 2011/2012 (McCollum et al 2015, doi: 10.4269/ajtmh.15-0095). These fragments represent only a small portion of the genome (5 genes), and as such, the samples’ exact positions on the phylogeny cannot be reliably reconstructed. With the available data, however, we can assert that two of the samples cluster with the recent MPXV sequences from Sud-Kivu (proposed Clade Ib), and, although divergent from the current outbreak, they likely represent some of the reservoir diversity present in the area surrounding the Kivu provinces (I14 and I37). The third sequenced sample from the 2011/2012 outbreak (I38) does not cluster with this diversity and lies in the distinct Clade Ia.

**Supplemental Figure 2.**
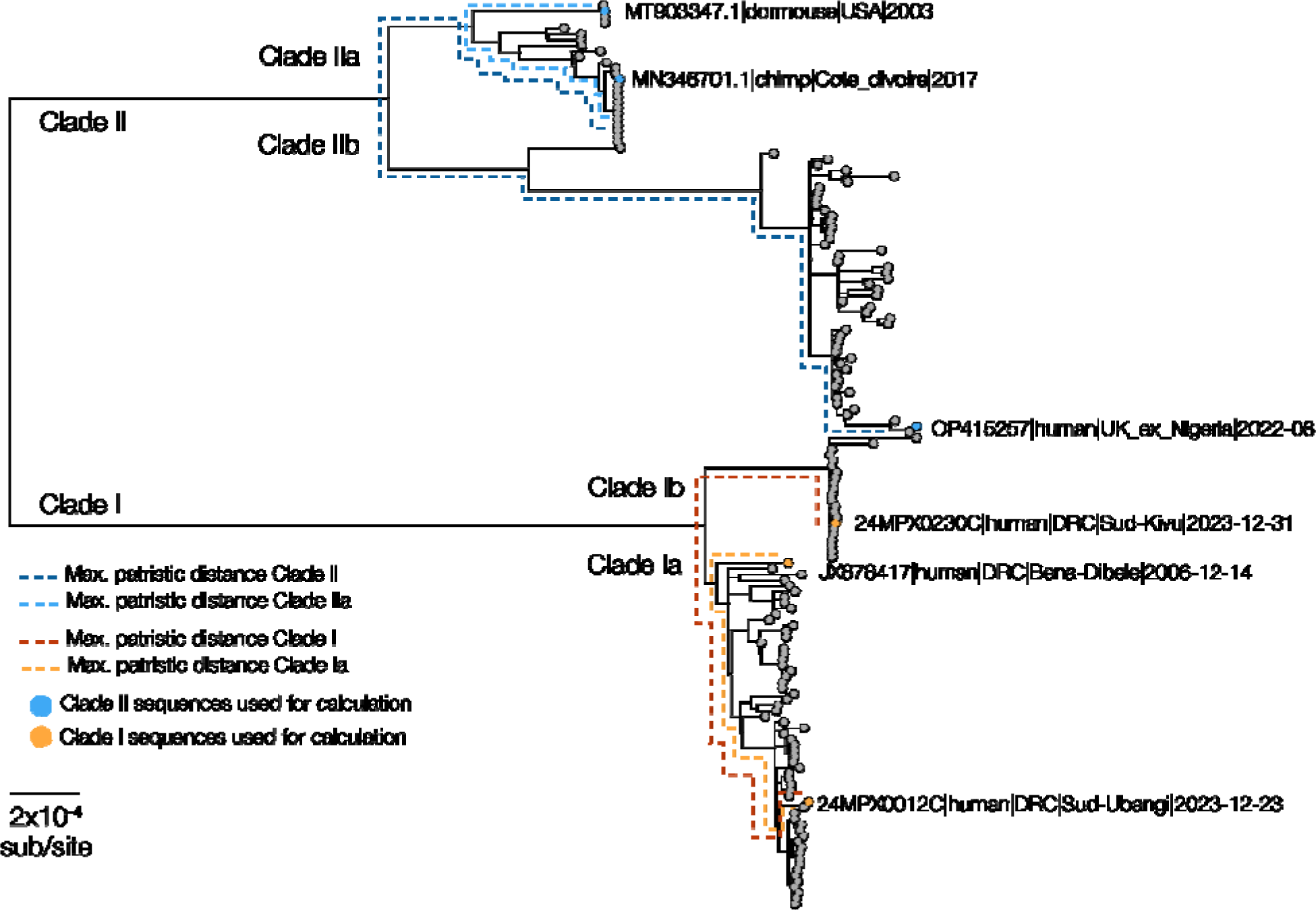
MPXV clades are indicated, including proposed Clade IIb, in the maximum likelihood phylogeny used to calculate maximum patristic distance and the increase in that with the addition of Clade IIb to Clade II and Clade IIb to Clade I. The sequences algorithmically identified for the calculation are indicated, and the backbone used for the pairwise calculations are marked with a dashed line.

**Supplemental Table 1.**
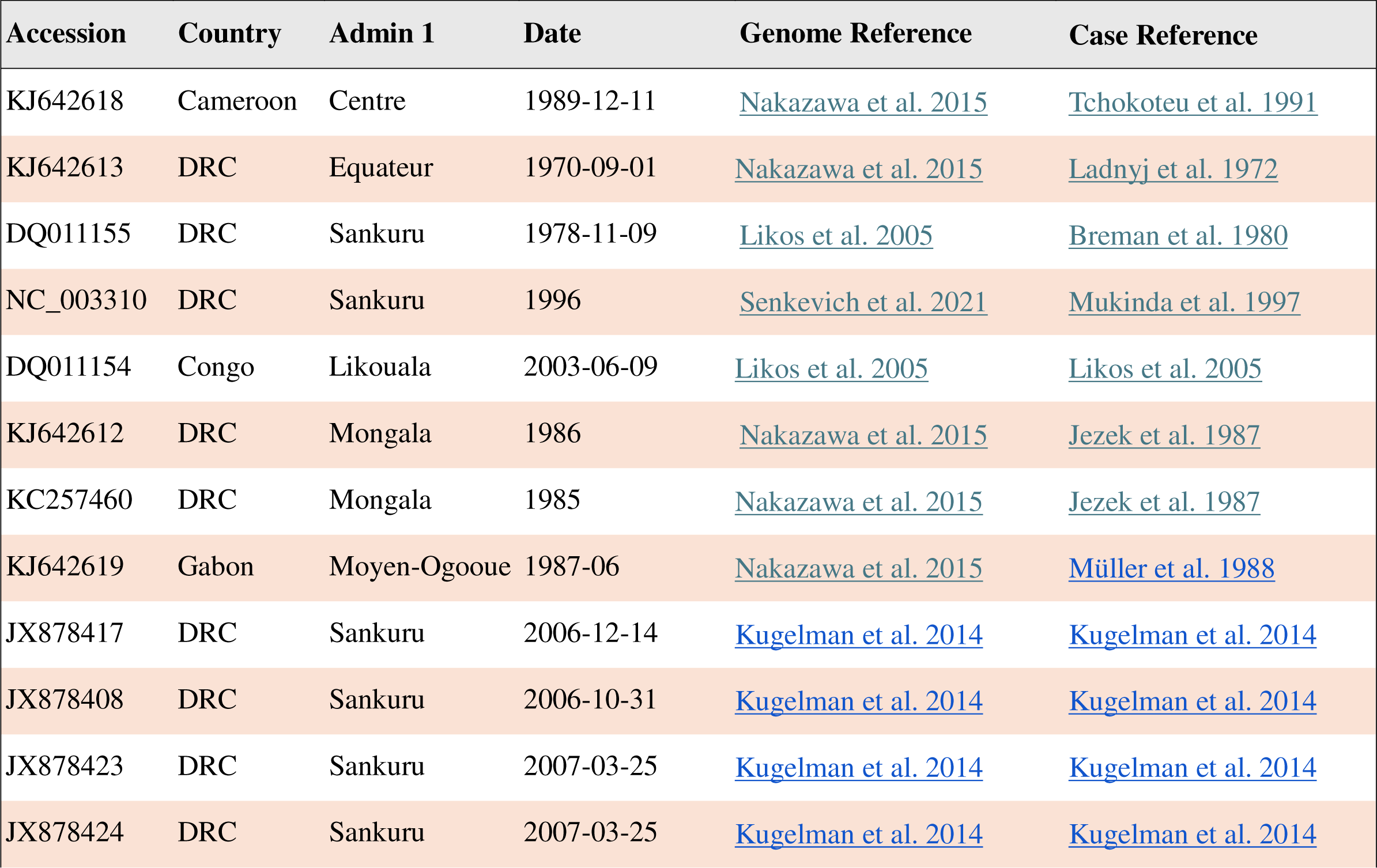
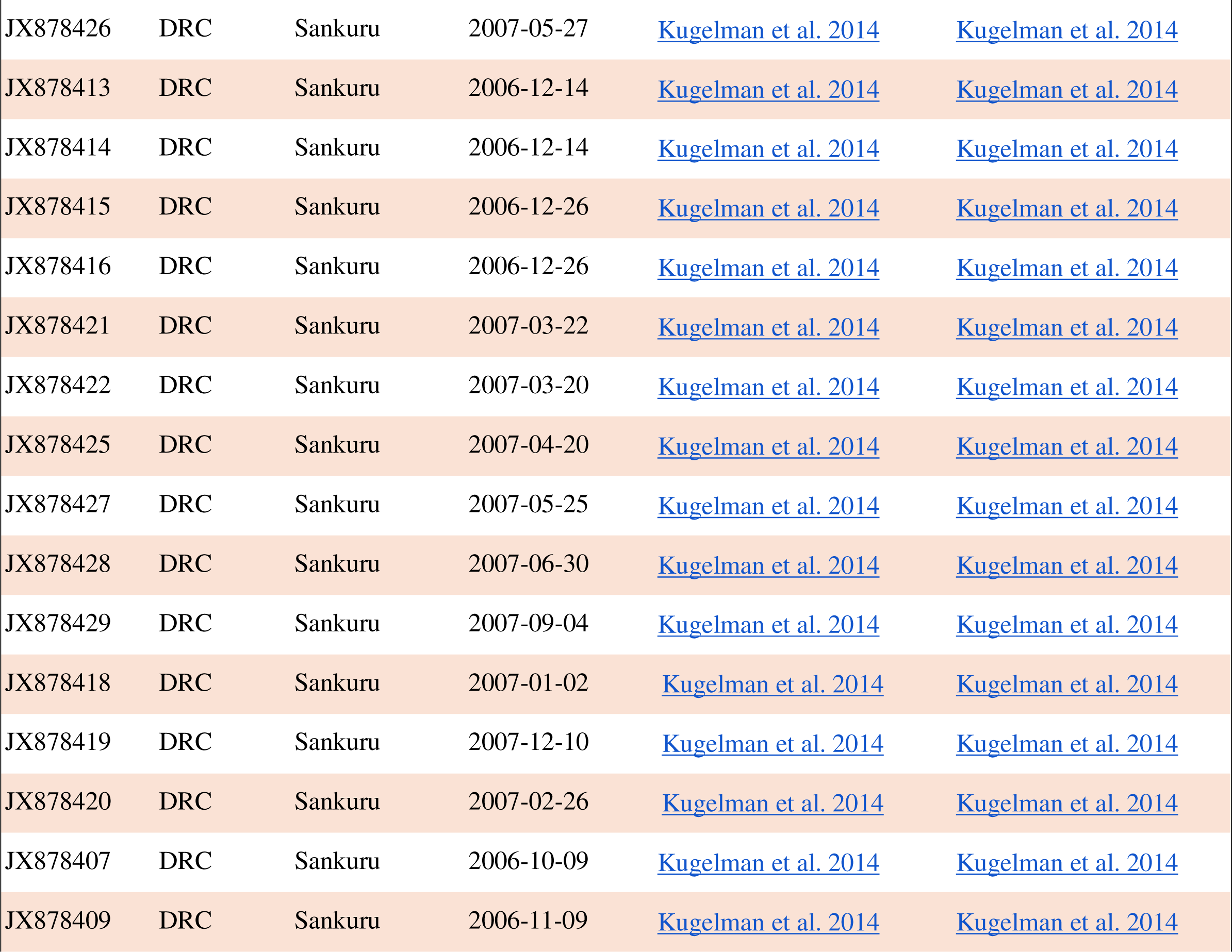
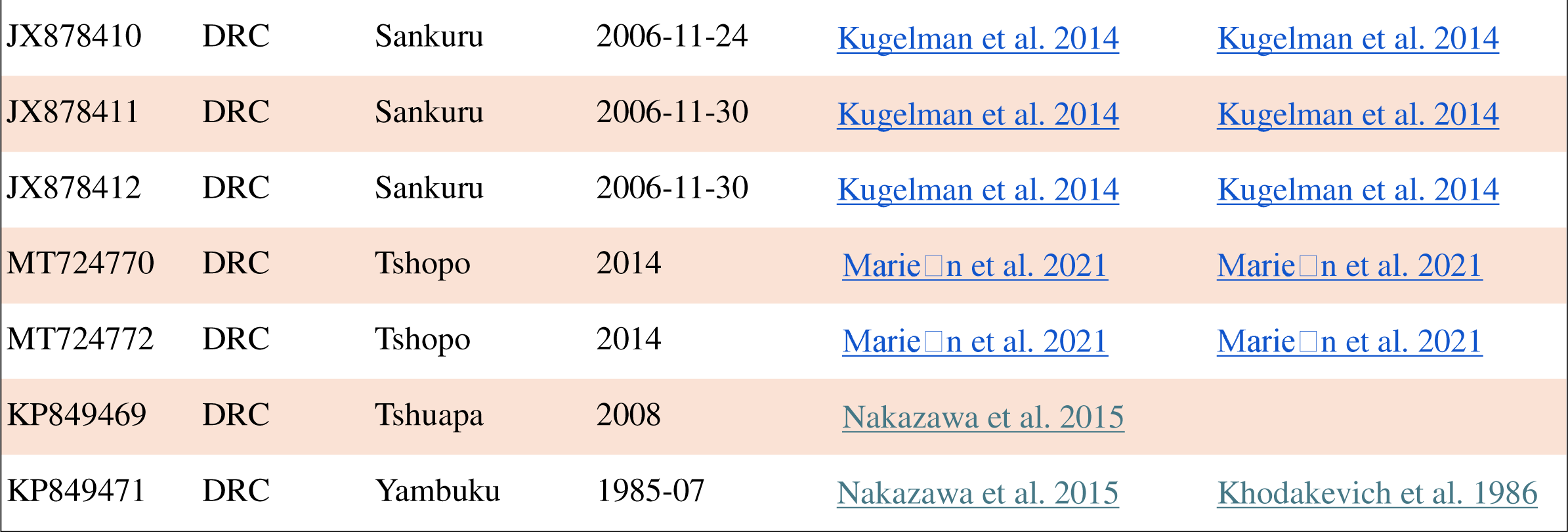
Previously published MPXV genomes used in this study with NCBI Genbank accession number (Accession), country of origin (Country), top level administration region (Admin 1), date of sample collection to highest known precision (Date), reference for the genome sequence (Genome Reference), and reference for case description were available (Case Reference).

**Supplemental Table 2.**
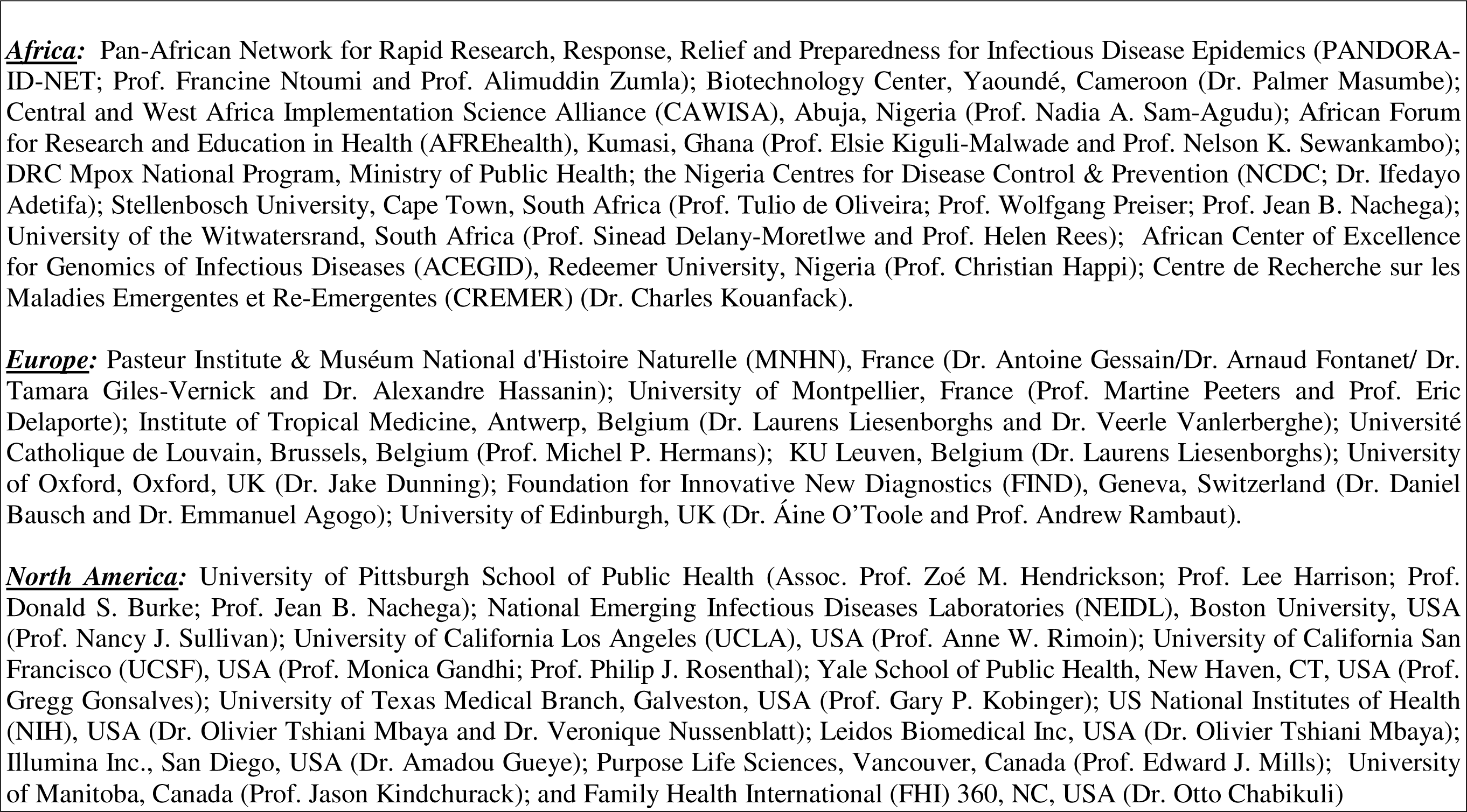
MpoxReC Collaborating Consortia, Institutions, and Organizations.

